# Risk factors for mortality of residents in nursing homes with Covid-19: a retrospective cohort study

**DOI:** 10.1101/2020.11.09.20228171

**Authors:** Clara Suñer, Dan Ouchi, Miquel Àngel Mas, Rosa Lopez Alarcon, Mireia Massot Mesquida, Eugènia Negredo, Núria Prat, Josep Maria Bonet-Simó, Ramon Miralles, Montserrat Teixidó Colet, Joaquim Verdaguer Puigvendrelló, Norma Henríquez, Michael Marks, Jordi Ara, Oriol Mitjà

**Author notes:** Corresponding author: Oriol Mitjà, Hospital Germans Trias i Pujol, Carretera Canyet s/n, 08916, Badalona, Spain.

## Abstract

**Background:** Nursing homes have shown remarkably high Covid-19 incidence and mortality. We aimed to explore the contribution of structural factors of nursing home facilities and the surrounding district to all-cause and Covid-19-related deaths during a SARS-CoV-2 outbreak.

**Methods:** In this retrospective cohort study, we investigated the risk factors of Covid-19 mortality at the facility level in nursing homes in Catalonia (North-East Spain). The investigated factors included characteristics of the residents (age, gender, comorbidities, and complexity and/or advanced disease), structural features of the nursing home (total number of residents, residents who return home during the pandemic, and capacity for pandemic response, based on an ad hoc score of availability of twelve essential items for implementing preventive measures), and sociodemographic profile of the catchment district (household income, population density, and population incidence of Covid-19). Study endpoints included all-cause death and Covid-19-related death (either PCR-confirmed or clinical suspicion).

**Findings:** The analysis included 167 nursing homes that provide long-term care to 8,716 residents. Between March 1 and June 1, 2020, 1,629 deaths were reported in these nursing homes; 1,089 (66□9%) of them were Covid-19-confirmed. The multivariable regression showed a higher risk of death associated with a higher percentage of complex patients (HR 1□09; 95%CI 1□05-1□12 per 10% increase) or those with advanced diseases (1□13; 1□07-1□19), lower capacity for implementing preventive measures (1□08; 1□05-1□10 per 1-point increase), and districts with a higher incidence of Covid-19 (2□98; 2□53-3□50 per 1000 cases/100,000 population increase). A higher population density of the catchment area was a protective factor (0□60; 0□50-0□72 per log10 people/Km^2^ increase).

**Interpretation:** Presence of residents with complex/advance disease, low capacity for pandemic response and location in areas with high incidence of Covid-19 are risk factors for Covid-19 mortality in nursing homes and may help policymakers to prioritize preventative interventions for pandemic containment.

**Funding:** Crowdfunding campaign YoMeCorono (https://www.yomecorono.com/), and Generalitat de Catalunya.

**Research in context:** *Evidence before this study:* We searched PubMed for studies exploring the management of Covid-19 in long-term care settings. The search was performed on May 1, 2020, and included the keywords “Covid-19”, “nursing home”, “long term care”, and “skilled nursing facility” with no language restriction. In addition to descriptive reports of Covid-19 mortality in the long-term care setting, we found studies providing evidence on the influence of age and comorbidities to mortality at the individual level. Some authors reported comparisons in the incidence and mortality of Covid-19 between facilities and country areas, and suggested the characteristics of each area/facility that may explain differences in mortality. However, we found no published works specifically investigating the contribution of structural features of the facility and sociodemographic characteristics of the area to explaining differences in Covid-19 mortality among long-term care facilities.

*Added value of this study:* This is the first analysis of risk of mortality at a facility level of residents with Covid-19 in nursing homes. We enrolled up to 167 nursing homes providing long-term care to 8,716 residents and we actively identified risk factors for Covid-19 mortality at the facility level. We found that nursing homes with lower capacity for pandemic response, and located in districts with a higher incidence of Covid-19 had significantly higher risks of Covid-19 mortality. The percentage of complex and/or advanced disease patients was also a risk factor.

*Implications of all the available evidence:* Our findings provide policymakers with critical information to prioritize long-term care facilities at higher risk when deploying preventative interventions to minimize mortality in this setting. The association between mortality within the nursing home and Covid-19 incidence in the catchment area reinforces the importance of preventing the entry of SARS-CoV-2 into facilities. Nursing homes with limited capacity to implement containment measures should be prioritized when deploying preventative interventions for minimizing Covid-19 mortality in long-term care facilities.

## Introduction

Six months after the first outbreak of the novel coronavirus disease (Covid-19), the global death toll associated with the pandemic amounted to nearly half a million.^1^ To date, various authors have reported on the major role of long-term care (LTC) facilities, such as nursing homes, in spreading SARS-CoV-2 to the most vulnerable populations during the Covid-19 pandemic.^2^ This group has experienced an extremely high death toll and also has overwhelmed local health systems. In some countries, LTC residents account for more than 50% of deaths attributed to Covid-19. In Catalonia (North-East Spain), the government reported approximately 1,810 deaths among residents of LTC facilities between March 15 and April 15.^3^

To date, large variations in Covid-19 death rates across LTC facilities have been observed. Whether the high death rates are linked to the structural features of such settings or the poorer health of individuals in these facilities compared to those living elsewhere is still unclear. Because of the different policy implications of the relative influence of these features, there is a need to deepen into the determinants of SARS-CoV-2 spread and mortality in LTC facilities.^4,5^

Potential risk factors of the residential setting are a communal living area, multiple residents in a single room, care provided by multiple caregivers – who may work across multiple different facilities, shortage of healthcare resources (e.g., tests, and personal protective equipment), limited access to skilled healthcare professionals, and the lack of specific guidance for managing the outbreak in the residential setting.^2,3,6^ In addition to these setting-specific risk factors, the higher death rates are likely associated primarily with older age, high levels of multi-comorbidity, disability, and immune senescence of old-age.^5,7–9^ Finally, some authors have identified risk factors associated with the characteristics of the population in the catchment area, such as the mean household income or the population density.^10,11^

We aimed to assess whether living in a nursing homes for LTC is associated with an increased risk of death from Covid-19 beyond the risk associated with age and chronic health conditions. We used data from nursing homes, including residents’ health characteristics, structural features, and the demographic and epidemiological profile of the district where the nursing home is located, to investigate the association between potential risk factors at the facility level and mortality in the residential setting during the SARS-CoV-2 outbreak in Spain.

## Methods

### Study setting and participants

This was a retrospective cohort study of Covid-19 mortality risk factors in the residential setting in Catalonia (North-East Spain). The study included clinical, mortality, and structural information corresponding to all public and private nursing homes in the administrative health region *Metropolitana Nord* (population 1,986,032 people) in Barcelona, Spain between March 1 and June 1, 2020, during the Covid-19 outbreak. Skilled nursing facilities (i.e., intermediate care) and mental health facilities were excluded from the analysis.

On March 1, 2020, the Department of Health of Catalonia launched a comprehensive disease control program to minimize Covid-19 spread and mortality among residents in nursing homes. The containment strategy was implemented in all LTC facilities in the study area and involved 64 primary care teams that reported daily information regarding the epidemiological status of each nursing home. The primary care teams provided preventive epidemiological recommendations, including the partition of communal living areas, isolation of suspected cases and contacts, guidance on personal protective measures to nursing home workers. In the advent of a confirmed or suspected case of Covid-19, the teams also conducted systematic screening of close contacts?or all residents, in centers with high incidence?using real-time reverse transcription–polymerase chain reaction (rt-PCR) from nasopharyngeal swabs.

The study protocol was approved by the institutional review board of Hospital Germans Trias Pujol.

### Data collection

Demographic and clinical data of residents were extracted from electronic medical records using a standardized data collection form.^12^ The structural and organizational features of each nursing home were gathered at facility assessment visits by the study team. The demographic and epidemiological profile of the nursing home district was retrieved from the Statistical Institute of Catalonia. Deaths were identified from the Mortality Registry of the Department of Health of Catalonia. All data were handled according to the General Data Protection Regulation 2016/679 on data protection and privacy for all individuals within the European Union and the local regulatory framework regarding data protection.

### Definitions

Variables regarding the residents’ health characteristics in each nursing home included demographic characteristics (i.e., age and gender), and clinical characteristics (i.e., number of comorbidities and percentage of residents with high dependence in activities of daily living, defined as a Barthel score < 50^13^). We also recorded the percentage of residents identified on electronic medical records as complex chronic patients (CCP) and patients with advanced chronic disease (ACD) by their primary care teams, according to clinical guidelines of the Catalan Health Department.^14^ These guidelines define CCP based on their clinical condition (e.g. multimorbidity, disability, difficult symptom control) and/or social environment (e.g., lack of support from family or caregivers, isolated household). Patients with ACD are those with advanced and irreversible chronic conditions that limit their life expectancy to approximately 12 months. Comorbidities were codified according to the ICD-10 system and included dementia, asthma or chronic obstructive pulmonary disease, hypertension, type-1 diabetes mellitus, type-2 diabetes mellitus, chronic kidney disease, cerebrovascular disease, cardiovascular disease.

Structural features of nursing homes were characterized according to their capacity for pandemic preparedness and response (SNQ12 score) and other relevant organizational variables such as current number of residents and percentage of residents who return home to live with their relatives due to the pandemic. The capacity of the nursing home for pandemic preparedness and response was assessed using an *ad hoc* set of 12 essential items that yields a score, called SNQ12 (*sine qua non* conditions for implementing the measures).^15^ The score indicates the number of unmet requirements, which ranges from 0 (all requirements are met) to 12 (all requirements are unmet). The requirements are related to three areas: 1) personal protective equipment (PPEs) (adequate supply, routine use, and use for waste management and cleaning/disinfection), 2) surveillance and communication (routine monitoring of symptom onset by non-healthcare professionals and communication of symptoms to occupational health services), and 3) cleaning and waste management (regular hand washing before and after contact with Covid-19 patients or their contacts, adequate laundry procedures, cleaning and disinfection of surfaces, use of an adequate disinfectant, adequate disposal of used PPEs).

The district demographic and epidemiological profile was assessed and defined using the household income and density of population in the municipality, and the population incidence of Covid-19 in the post code district (lowest administrative division) where the nursing home is located.

Deaths were classified as Covid-19-related when individuals had a positive rt-PCR or a clinical suspicion of Covid-19. Clinical suspicion of Covid-19 was defined based on the national guidelines available at the time as individuals with clinical features of acute respiratory disease of sudden onset and any severity, primarily characterized by fever, cough, and shortness of breath. Other symptoms such as odynophagia, anosmia, dysgeusia, muscular pain, diarrhea, chest pain, or headache could also be considered suggestive of SARS-CoV-2 at the physician’s discretion.

### Statistical Analysis

Continuous and categorical variables were presented as the mean and standard deviation (SD) (or median and interquartile range [IQR], defined by 25^th^ and 75^th^ percentiles) and number (%), respectively. The excess deaths were defined as the difference between deaths reported in 2020 and the median of 2016-2019 for the same months of the year; the Covid-19 contribution to the excess deaths was computed by the difference between confirmed or suspected Covid-19 deaths and all-cause mortality. In our primary analysis to determine the risk factors associated with mortality, we used univariate and multivariate Poisson regression models at facility level. Variables for the multivariate model were treated as linear, and were chosen using an Akaike Information Criteria (AIC)-based backward stepwise procedure. Results were presented as the hazard ratio (HR) and the 95% confidence interval (CI). In a secondary analysis, we grouped the nursing homes according to their characteristics using cluster analysis based on k-nearest neighbor classifier.^16,17^ The resulting clusters were described in a heatmap that represents the intensity of each characteristic based on the difference (below or above) between the average of the given cluster and that of the overall sample. We used a random forest classifier and the Gini measure of importance^18^ to determine the weight of each variable in each cluster. The significance threshold was set at a two-sided alpha value of 0.05. All analyses and plots were performed using R version 3.6.^19^

## Results

### Characteristics of the nursing homes

The analysis included 167 nursing homes providing long-term care to 8,716 residents. Table 1 summarizes the characteristics of the nursing homes included in the analysis. The mean age was 87□1 years, 56□6% of them were classified as CCPs and/or ACD patients, and 82□1% were identified as highly dependent. The median SNQ12 score was 1□4 unmet preventative items, reflecting an overall high level of pandemic preparedness. The individual demographic, clinical, and epidemiological characteristics of included residents are summarized in Table S1.

### Mortality

Between March 1 and June 1, 2020, a total of 1,629 deaths were reported in the nursing homes included in the analysis. Of these, 1,089 (66□9%) were registered as Covid-19 deaths in the mortality registry of the Department of Health. The cause of the death of the remaining 671 deaths could not be confirmed. Overall, the excess deaths in the analyzed nursing homes compared with the same period in the four previous years were estimated to be 971 deaths; Covid-19-confirmed deaths accounted for 89□2% of all excess mortality (Figure 1). At the nursing home level, the median (IQR) mortality rate was 14□3 (7□6 – 26□1) deaths/100 residents/3-month study period for all-cause death, and 3□9 (0.0 – 18□4) for Covid-19 confirmed deaths.

### Risk factors for mortality

According to the multivariate analysis, the risk of Covid-19 related deaths was higher in nursing homes with a higher percentage of CCP patients (hazard ratio 1□09; 95%CI 1□05-1□12 per 10 units increase) or ACD patients (1□13; 1□07-1□19 per 10 units increase), lower capacity for pandemic preparedness and response (1□08, 1□05-1□10 per unit increase) and located in areas with high incidence of Covid-19 (2□98; 2□53-3□50 per 1000 cases/100,000 population increase) (Table 2). The risk factors of all-cause death were the same as those of Covid-19 related death. For Covid-19-related deaths, the univariate analysis revealed a higher risk of death in nursing homes with a high percentage of residents who returned home to live with their relatives. This variable was selected in the stepwise method, but it was not significantly associated with Covid-19-related deaths in the multivariate analysis. The only variable associated with lower all-cause deaths was living in high-density population areas (0□60; 0□50-0□72 per log 10 people/km2). The risk factors significantly associated to all-cause death were the same as those of Covid-19 related death (Table 2).

### Comparison of characteristics among phenogroups

The cluster analysis based on the k-nearest neighbor classifier identified eight groups of nursing homes that were significantly different from each other. Resident health characteristics, structural features, and sociodemographic factors were stratified according to each phenogroup. Figure 2 illustrates the intensity of each characteristic (i.e., the extent of the difference between the mean of a given cluster and that of the entire sample) in the resulting clusters and the contribution of each characteristic to the definition of a given phenogroup. Key characteristics of each cluster were as follows:

Nursing homes in **cluster 1** were placed in low densely populated areas with high population incidence of Covid-19, and high household income; **cluster 2** were facilities with a high proportion of CCP and ACD patients, and located in areas with low population incidence of Covid-19; **cluster 3** had low proportion of CCPs and highly dependent residents; **cluster 4** had higher number of residents than the median, although with a very low proportion of CCPs; nursing homes in this cluster were placed in areas with low household income; **cluster 5** had low proportion of ACD patients and dependent residents, and had higher number of residents that returned home with their relatives; **cluster 6** were placed in areas with high household income and low population incidence of Covid-19; **cluster 7** had high proportion of CCP and ACD patients; nursing homes in this cluster were located in densely populated areas; **cluster 8** had a high SNQ12 score?indicating very limited capacity for pandemic preparedness and response?and high proportion of CCPs and older residents than the median.

### Association of phenogroups with mortality

The mortality rate in each phenogroup is shown in Figure 3. During the study period, the median (IQR) proportion of all-cause deaths and Covid-19-related deaths in the eight nursing home clusters was 12□3% (7□6 - 26□1) and 3□9% (0□0 - 18□4), respectively. Clusters 1, 4, and 8 had a greater all-cause mortality rate than the median. Correspondingly, clusters 1, 7, and 8 had a greater Covid-19-related mortality rate than the median.

## Discussion

To our knowledge this is the first study on risk of mortality at a facility level of residents with Covid-19 in nursing homes. Our analysis revealed that a ten percent increase in the proportion of residents with complex or advanced diseases increased the mortality risk by 9% and 13%, respectively; a 1-point increase in the 12-points score of unmeet measures for containing SARS-CoV-2 spread increases the mortality risk by 8%, and an increase in population incidence of 1000 Covid-19 cases per 100,000 population increases the mortality risk by 198%. Location in a highly densely populated area was the only factor associated with a reduced mortality rate, which might be related to improved access to hospitals with intensive care units in urban areas, as previously suggested.^20^

The clustering of nursing homes according to their residents’ profile and structural capacities provided a global perspective of the type of nursing homes that might be more susceptible to Covid-19 mortality in the advent of future outbreaks. Consistently with our regression analysis, clusters with greater mortality than the median (phenogroup numbers 1,4,7, and 8) were all located in neighborhoods with high incidence of Covid-19. These results align with previous studies that reported a significant relationship between LTC mortality and Covid-19 incidence in the catchment area.^5,10,11^ The increasing evidence on the influence of the local incidence of Covid-19 on mortality underscores the paramount importance of early detection?and response to?SARS-CoV-2 entry into facilities?often with new residents, staff, or visitors?for preventing uncontrolled outbreaks in this setting.^5,21^ These finding also suggests that population efforts to contain Covid-19 incidence may also contribute to reducing Covid-19 deaths at their local nursing homes

The multiple regression and cluster analysis were also consistent regarding the importance of the capacity of the nursing home for pandemic preparedness and response. Although most nursing homes showed low SNQ12 scores?indicating few unmet needs for applying containment measures?facilities in phenogroup 8, characterized by higher SNQ12 scores (mean of 5 unmet items over a total of 12 essential requirements) than the median, experienced high mortality levels.

Finally, our multivariate analysis revealed a significant relationship between higher percentages of CCP and/or ACD patients and increased mortality risk. According to local clinical guidelines, CCP and ACD patients are characterized by high clinical complexity and the presence of an advanced?often terminal?disease, respectively,^14^ suggesting an increased likelihood of death in the advent of any infection or acquired disease. Interestingly, phenogroup no. 2, characterized by the higher health risk of its residents, had similar mortality than phenogroups 4, 5, and 6, with a more favorable resident health profile. These conflicting results suggest that the mechanisms driving mortality risk in nursing homes are complex and may depend on the conjunction of various factors.

Our analysis had the intrinsic limitations of retrospective studies, particularly regarding data completeness. Owing to the overload of the healthcare system during the investigated period, a large number of deaths could not be tested for SARS-CoV-2 PCR and remained unconfirmed. We were unable to gather information regarding the worker profiles in each nursing home. Unlike skilled nursing homes aimed at intermediate care or mental health resources, which tend to be coordinated by the healthcare authorities, non-specialized nursing homes aimed at long-term stay are a case-mix of organizational models. Hence, the inclusion of the characteristics of the work team profile (e.g., skills, resident/worker ratio, and presence of physicians) might have provided interesting insights regarding the capacity of the residence to cope with the outbreak.^22^

Our results raise important policy implications by suggesting structural factors of the nursing homes and their surrounding districts that are important drivers of Covid-19-related mortality in this setting. Identification of facilities with low capacity for pandemic response, located in areas with high incidence of Covid-19 and low density of population (e.g., rural areas) could help public health officers to identify facilities where preventative interventions need to be prioritized. The presence of complex patients and those with advanced chronic diseases also increased mortality risk, though these factors alone seem not to explain mortality trends at facility level. Efforts should be geared to protecting older adults living in the highest risk facilities.

## Data Availability

Data referred to in the manuscript are available upon request to the corresponsding autohr

## Contributors

CS, DO, MAM, RLA and OM designed the study. DO, MAM, RLA, EN, MMM, NP, JMB-S, RM, MTC, JVP, NH, JA collected the data. CS, DO, MM, OM analyzed the data. CS, DO, OM interpreted the data. CS, DO, OM wrote the manuscript. All authors reviewed and approved the final version of the manuscript

## Acknowledgments

The authors would like to thank Gerard Carot-Sans for providing medical writing support during the preparation of the manuscript.

---

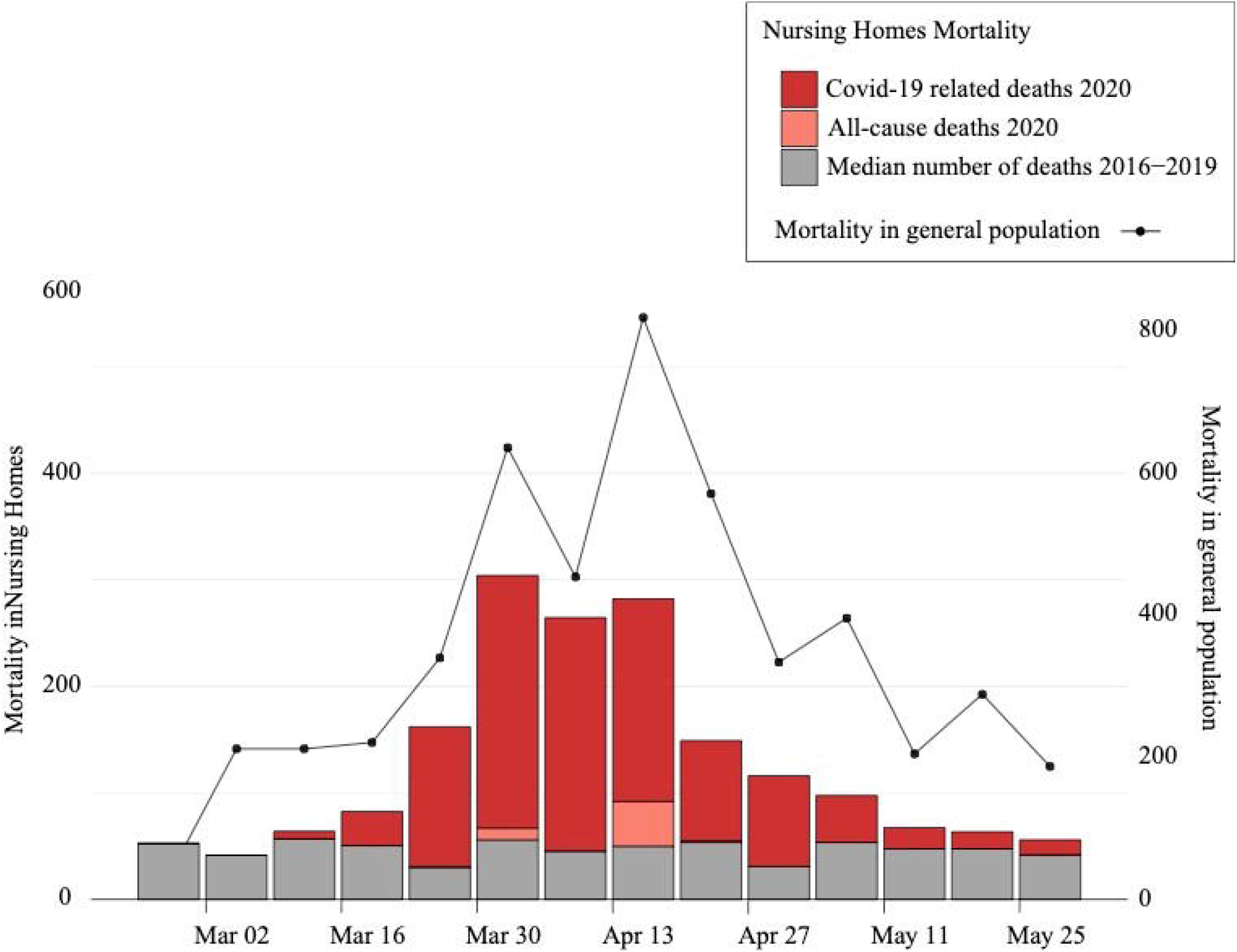

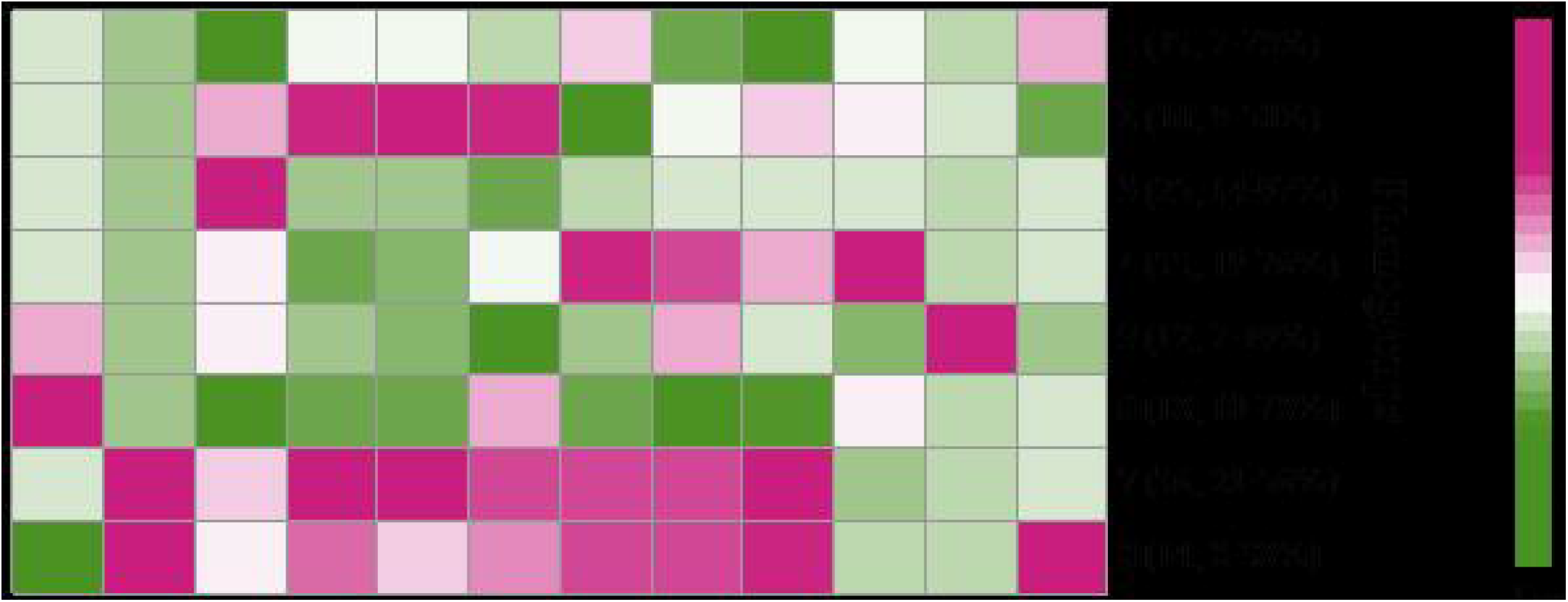

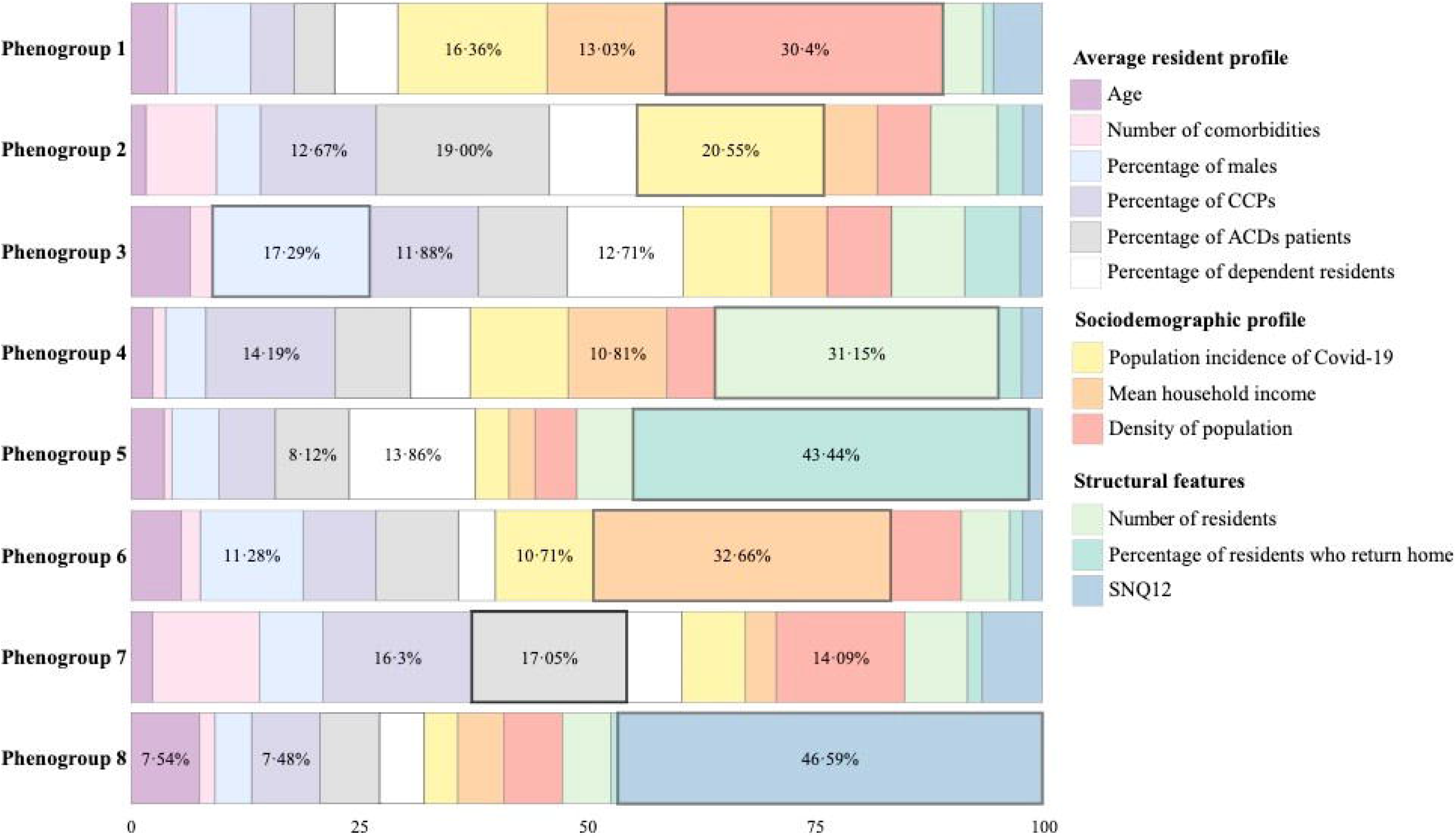

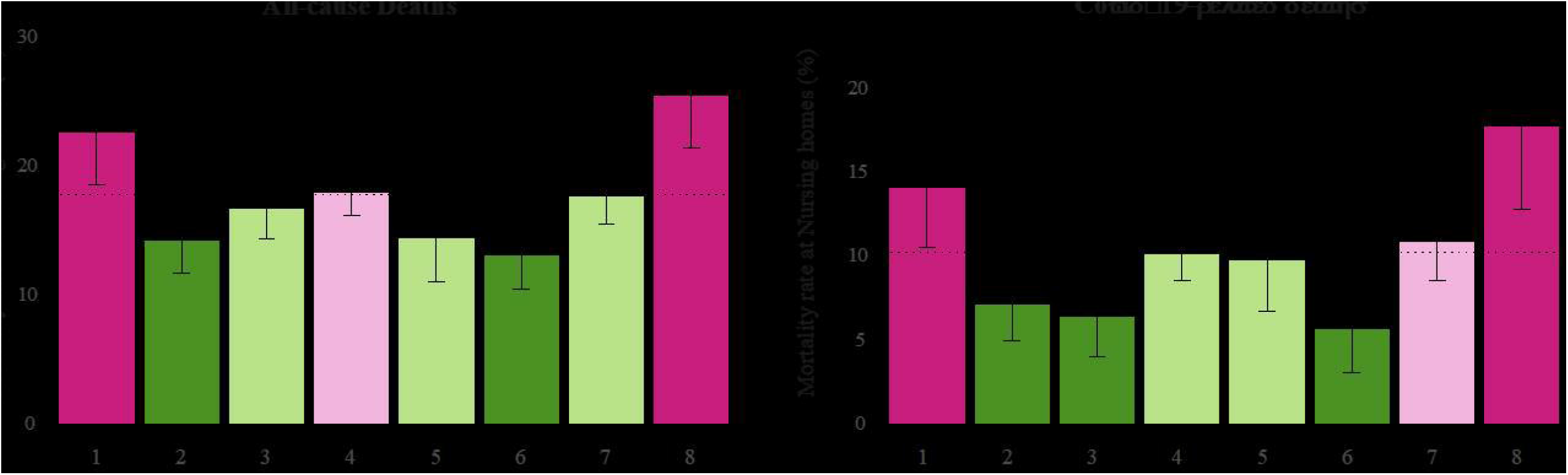

