## Supplementary figures and images for "Risk factors for mortality of residents in nursing homes with Covid-19: a retrospective cohort study"

### Supplementary Figure 1

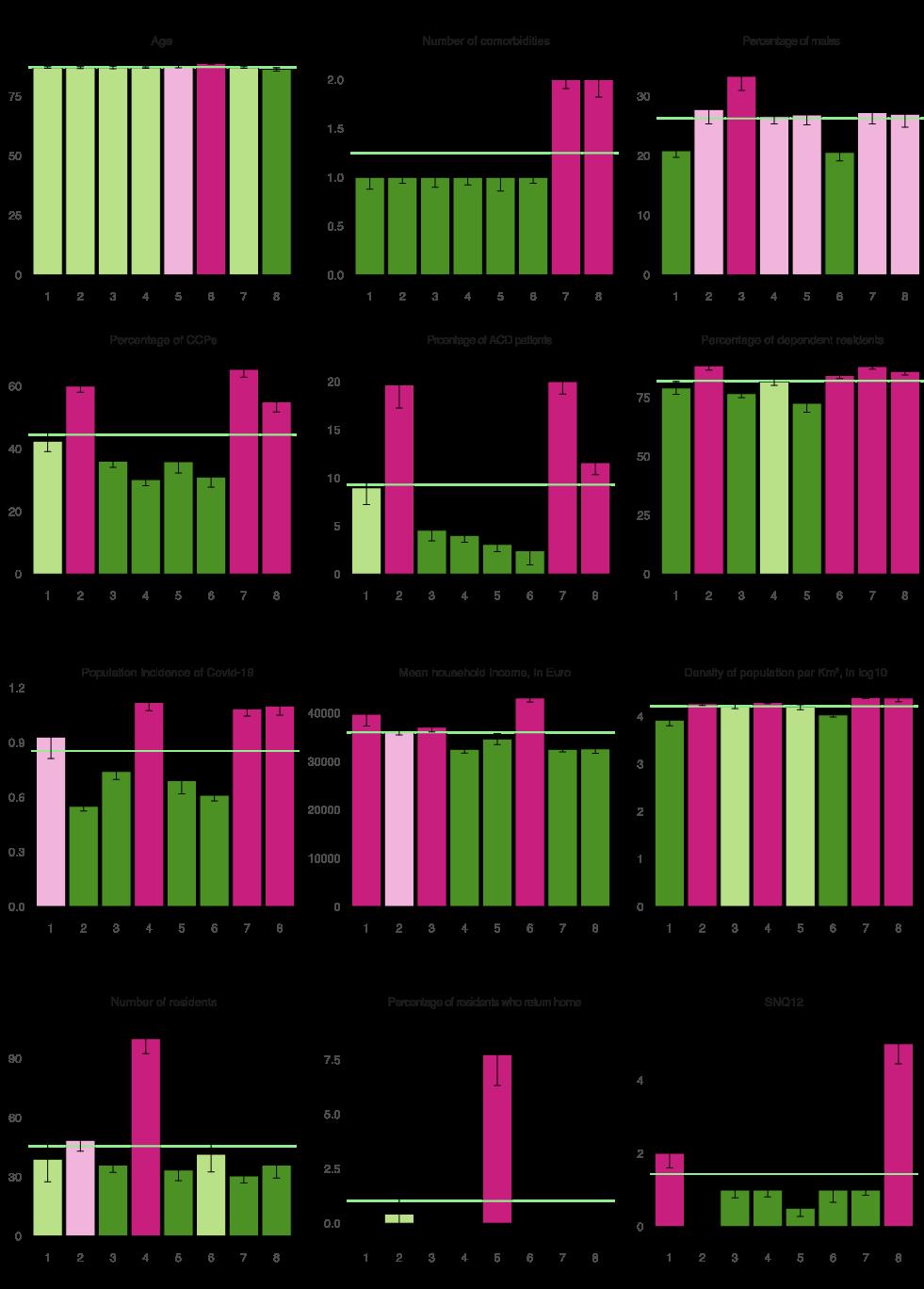
