## Supplementary material for "Risk factors for mortality of residents in nursing homes with Covid-19: a retrospective cohort study": Tables and Figures

| **Table 1. Characteristics of nursing homes** | |
| --- | --- |
|  | **Total** |
|  | **(N = 167)** |
| **Average resident profile** |  |
| Age of residents, in years | 87·1 (2·1) |
| Percentage of male residents | 26·4 (9·6) |
| Number of comorbidities | 1·5 (0·6) |
| Percentage of CCPs | 46·1 (17·3) |
| Percentage of ACD patients | 10·5 (8·9) |
| Percentage of dependent residents* | 82·1 (9·5) |
| **Structural features** |  |
| SNQ12 score | 1·4 (1·7) |
| Current number of residents | 46·2 (29·8) |
| Percentage of residents who return home | 1·4 (3·0) |
| **District demographic and epidemiological profile** |  |
| Mean household income, in Euro | 36099·6 (5527·5) |
| Density of population per km2, in log10 | 17·9 (9·5) |
| Population incidence of Covid-19 | 0·9 (0·3) |

Data are mean (SD). CCP=complex chronic patient. ACD=advanced chronic disease. SNQ12=number of unmet epidemic and infection control preparedness requirements (0-12). *Barthel score < 50.

**Figure 1. Excess mortality of 2020 relative to the average of the past four years (2016-2019)**

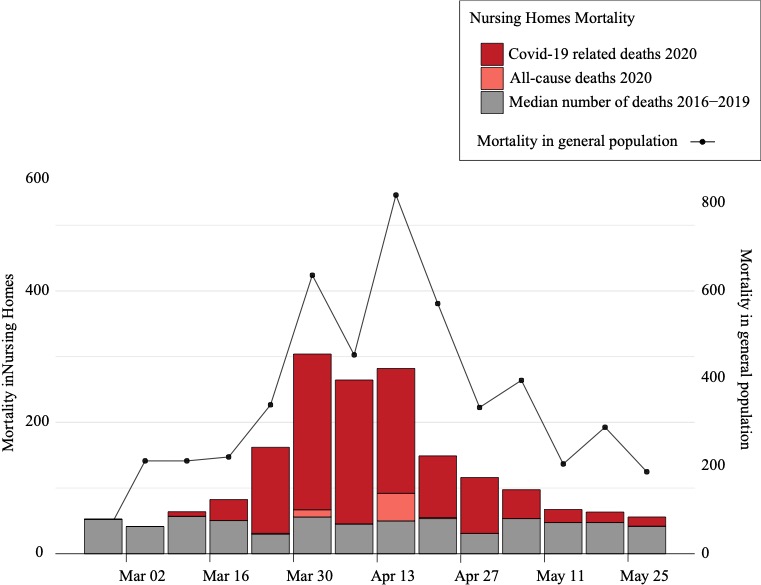

Bars show the number of weekly deaths reported in 2020 in all nursing homes included in the analysis over the study period. Deaths reported in 2020 have been classified as Covid-19 confirmed and unconfirmed, which include deaths of individuals with suspected Covid-19 diagnosis. The median number of deaths for the same weeks in the previous 4 years (2016-2019) is shown in grey. The continuous line shows the death toll attributed to Covid-19 in the general population of the catchment area.

**Table 2. Estimated effect of long-term care facilities’ features in all deaths and Covid-19 related deaths.**

|  | **All-cause deaths** | | | **Covid-19-related deaths** | | |
| --- | --- | --- | --- | --- | --- | --- |
|  | **Univariate analysis** | **Multivariate analysis** | | **Univariate analysis** | **Multivariate analysis** | |
| **Average resident profile** | **Hazard Ratio (95% CI)** | **Hazard Ratio (95% CI)** | **p-value** | **Hazard Ratio (95% CI)** | **Hazard Ratio (95% CI)** | **p-value** |
| Age of residents in each facility § | 0·99  (0·83-1·18) | ·· |  | 0·99  (0·97-1·01) | ·· |  |
| Percentage of male residents | 1·01  (1·03-1·05) | ·· |  | 1·00  (0·95-1·05) | ·· |  |
| Number of comorbidities § | 1·15  (1·08-1·22)* | ·· |  | 1·35  (1·25-1·46)* | ·· |  |
| Percentage of CCPs | 1·04  (1·02-1·06)* | 1·04  (1·02-1·06) | 0·0015 | 1·10  (1·06-1·11)* | 1·09  (1·05-1·12) | <0·0001 |
| Percentage of ACD patients | 1·09  (1·05-1·13)* | 1·09  (1·04-1·13) | 0·0002 | 1·15  (1·10-1·20)* | 1·13  (1·07-1·19) | <0·0001 |
| Percentage of dependent residents | 1·03  (1·01-1·07) | ·· | ·· | 1·00  (0·95-1·05) | ·· | ·· |
| **Structural features** |  |  |  |  |  |  |
| SNQ12 § | 1·06  (1·04-1·08)* | 1·04  (1·03-1·07) | <0·0001 | 1·11  (1·09-1·14)* | 1·08  (1·05-1·10) | <0·0001 |
| Current number of residents § | 1·00  (0·99-1·01) | ·· | ·· | 1·00  (0·98-1·02) | ·· | ·· |
| Percentage of residents who return home | 1·03  (0·91-1·14) | ·· | ·· | 1·17  (1·01-1·31)* | ·· | ·· |
| **Sociodemographic profile** |  |  |  |  |  |  |
| Mean household income,  in Euro† | 0·95  (0·90-1·02) | ·· | ·· | 0·95  (0·86-1·04) | ·· | ·· |
| Density of population, log10 people/km^2^ ¶ | 0·84  (0·74-0·95)* | 0·67  (0·59-0·77) | <0·0001 | 0·91  (0·77-1·08) | 0·60  (0·50-0·72) | <0·0001 |
| Population incidence of  Covid-19‡ | 1·67  (1·48-1·87)* | 1·79  (1·59-2·03)* | <0·0001 | 2·72  (2·33-3·18)* | 2·98  (2·53-3·50)* | <0·0001 |

CCP= complex chronic patient; ACD= advanced chronic disease; Dependent resident=Barthel score < 50; SNQ12= number of unmet essential items for implementing preventive measures (0-12).

Hazar ratios and 95% CI are shown. All continuous predictors are mean-centered and scaled by 1 standard deviation.

HR represents the estimated effect for an increase of 10 units, unless otherwise indicated.

§ HR for an increase in 1 unit.

† HR for an increase of 10,000€/annum in mean household income.

¶ HR for an increase in 1 log10 people/km^2^.

‡ HR for an increase in incidence of Covid-19 of 1,000 cases/100,000 population.

*p-value for univariate analysis <0·05

**Figure 2.** Heatmap of nursing home phenogroups.

**A**

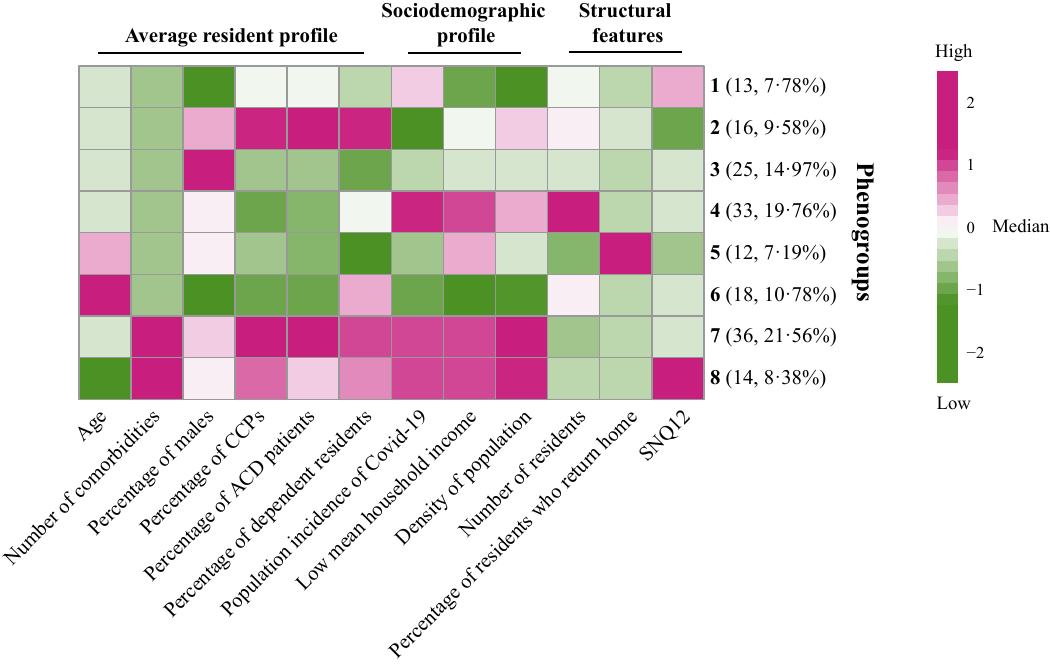

**B**

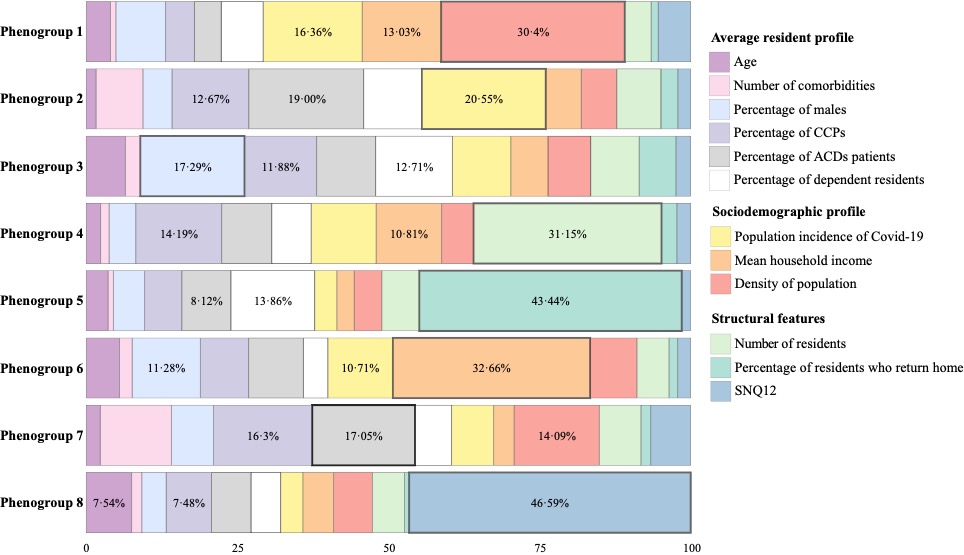

**A) Heatmap of nursing home phenogroups**. For each characteristic (x-axis), the extent of the difference between the mean of a given phenogroup and the median of the entire sample is illustrated with the following color code: green tones indicate a mean of the phenogroup below the median of the entire sample, whereas purple tones indicate a mean of the phenogroup above the median of the entire sample. In both cases, more intense colors represent greater differences between the phenogroup and the whole sample. For each phenogroup, the number of nursing homes is indicated (n, % of total). Figure S1 (Supplementary material) provides further details regarding the mean (SD) of the characteristics in each phenogroup. **B) Contribution of variables to each phenogroup.** Percentage is shown for the three variables with greater weight. Black squares highlight the most important variable.

**Figure 3. Mortality at the phenogroup level for all-cause deaths and Covid-19 related deaths.**

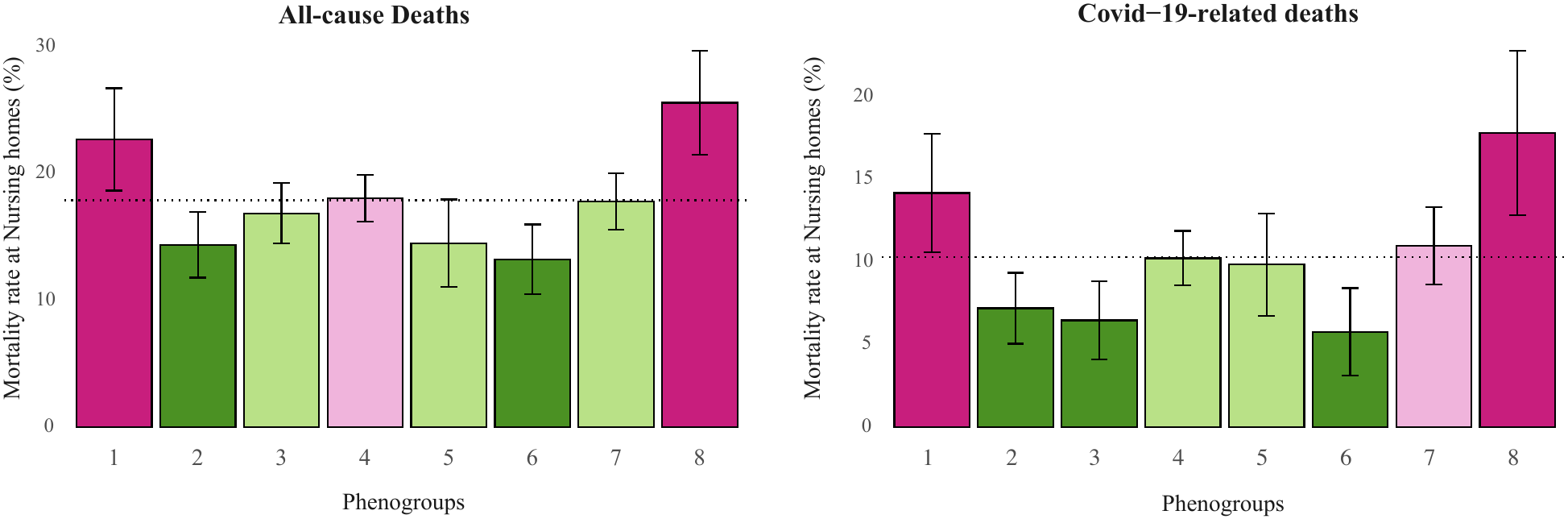

The dotted line shows the median mortality rate for the entire sample. Bars show the mean mortality rate of each phenogroup; error lines represent the standard error of the mean (SEM). Red and green indicate mean phenogroup mortality higher and lower than the total median, respectively. For both colors, light tones indicate that the SD of the phenogroup encompasses the overall median, whereas intense tones indicate that the whole SD range is above (red) or below (green) the total median.

| **Supplementary Table 1. Demographic and clinical characteristics of residents included in the analysis** | |
| --- | --- |
|  | Total (N=8·716) |
| **Demographics** |  |
| Age in years, mean *(SD)* | 85·7 (9·0) |
| Males | 2333 (26·8) |
| **Clinical Characteristics** |  |
| **Comorbidities** |  |
| Number of selected comorbidities, *n (%)* |  |
| 0 | 2142 (24·6) |
| 1 | 2528 (29·0) |
| 2 | 1869 (21·4) |
| >2 | 2177 (25·0) |
| Type of Comorbidity* *n (%)* |  |
| Dementia | 3954 (45·4) |
| COPD or asthma | 885 (10·2) |
| Hypertension | 4195 (48·1) |
| Diabetes mellitus type 1 | 19 (0·2) |
| Diabetes mellitus type 2 | 1704 (19·6) |
| Chronic kidney disease | 1707 (19·6) |
| Cerebrovascular diseases | 231 (2·7) |
| Cardiovascular diseases | 1228 (14·1) |
| **Chronic patient profile,** *n (%)* |  |
| Complex Chronic Patients (CCPs) | 3703 (42·4) |
| Advanced Chronic Disease (ACD) patients | 752 (8·6) |
| **Medication** |  |
| Polypharmacy (≥5 drugs), *n (%)* | 7305 (83·8) |
| Prescription of drugs of interest†, *n (%)* |  |
| Angiotensin-converting-enzyme inhibitors | 2184 (25·1) |
| Antiplatelets | 2602 (29·9) |
| Anticoagulants | 1146 (13·1) |
| Inhibitors of the sodium-glucose transport protein 2 | 16 (0·2) |
| Angiotensin II receptor blockers | 1098 (12·6) |
| Oral Corticoids | 220 (2·5) |
| Clozapine | 17 (0·2) |

Data are n (%) unless otherwise specified. *Categories are not mutually exclusive. †Drugs of interest are those with potential effect on the course of Covid-19 disease.

**Supplementary Figure 1. Nursing homes characteristics for each phenogroup.**

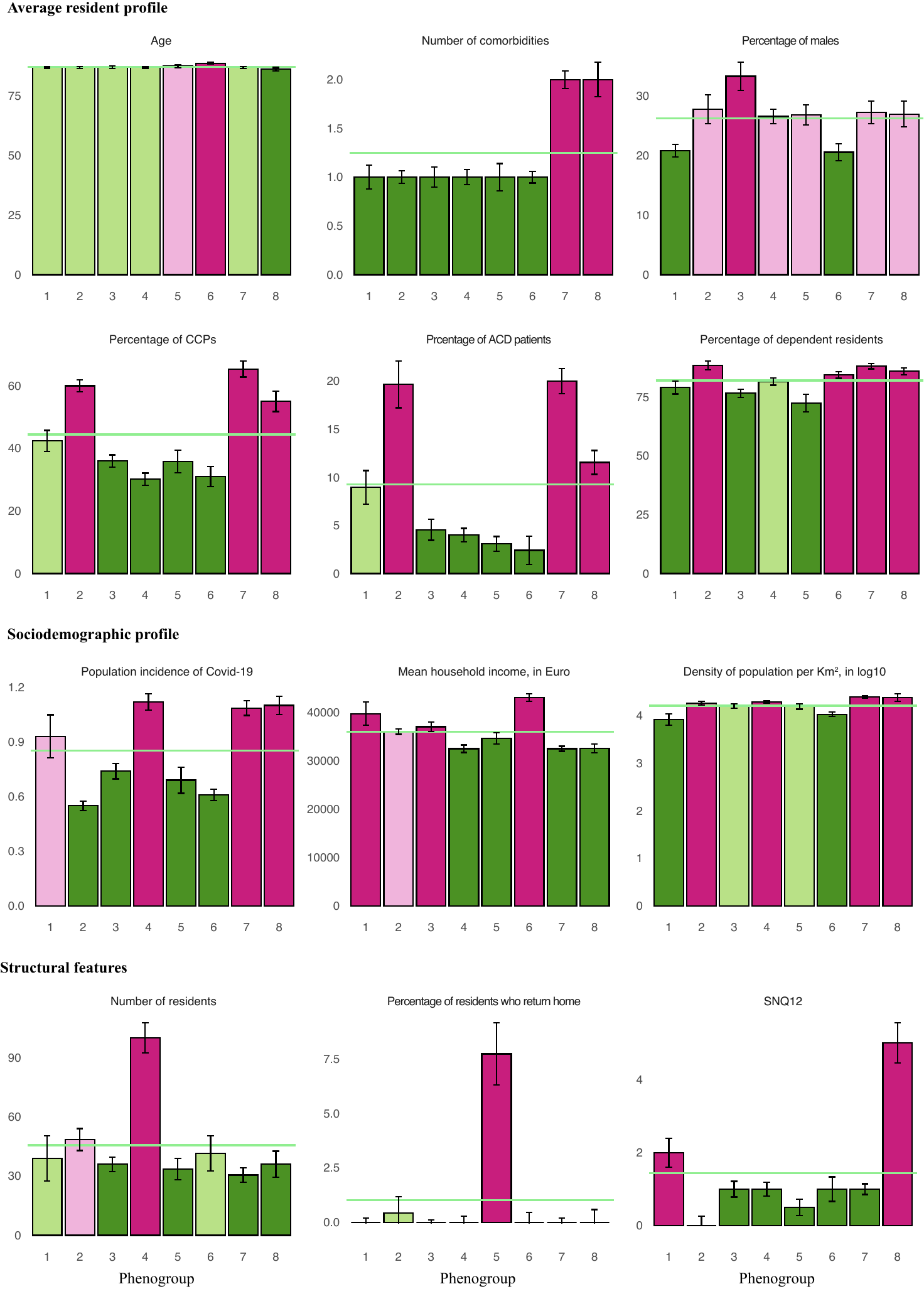

**Supplementary Figure 1.** For each phenogroup (x-axis), barplots show the mean (SD) of a given characteristic. The median of the entire sample is shown with a green line. The extent of the difference between the mean of a given phenogroup and the median of the entire sample is illustrated with the following color code: green tones indicate a mean of the phenogroup below the median of the entire sample, whereas purple tones indicate a mean of the phenogroup above the median of the entire sample. In both cases, more intense colors represent greater differences between the phenogroup and the whole sample.
